# The efficacy and safety of bamlanivimab treatment against COVID-19: A meta-analysis

**DOI:** 10.1101/2021.08.24.21262580

**Authors:** Huai-rong Xiang, Bei He, Yun Li, Xuan Cheng, Qi-zhi Zhang, Wen-xing Peng

**Author notes:** **Corresponding author:**, Email address, Full postal address: Department of Pharmacy, the Second Xiangya Hospital, Central South University, #139, Middle Renmin Road, Changsha, Hunan 410011, P.R. China.

## Abstract

**Background:** Bamlanivimab is routinely used in the treatment of coronavirus disease 2019 (COVID-19) in worldwide. We performed a meta-analysis to investigate the efficacy and safety of bamlanivimab treatment in patients with COVID-19.

**Methods:** We searched articles from Web of Science, PubMed, Embase, the Cochrane Library and MedRxiv between 30 January 2020 and August 5, 2021. We selected randomized clinical trials (RCTs) and observational studies with a control group to assess the efficiency of bamlanivimab in treating patients with COVID-19.

**Results:** Our meta-analysis retrieved 3 RCTs and 7 cohort studies including 14461 patients. Bmlanivimab may help outpatients to prevent hospitalization or emergency department visit (RR 0.41 95%CI 0.29 to 0.58), reduce ICU admission (RR 0.47 95%CI 0.23 to 0.92) and mortality (RR 0.32 95%CI 0.13 to 0.77) from the disease. The combination of bamlanivimab and etesevimab may had a greater potential for positive treatment outcome.

**Conclusion:** Bamlanivimab has demonstrated clinical efficacy on mild or moderate ill patients with COVID-19 to prevent hospitalization, reduce severity and mortality from the disease. Combinations of two or more monoclonal antibody increase the effect. Well-designed clinical trials to identify the clinical and biochemical characteristics in COVID-19 patients’ population that could benefit from bamlanivimab are warranted in the future.

**Key points:** *Question:* Can bamlanivimab treat COVID-19 patients? What are the factors that have great impact on the treatment outcome?

*Findings:* In this meta-analysis that retrieved 3 RCTs and 7 cohort studies and included 14461 adults, Bmlanivimab may help outpatients to prevent hospitalization or emergency department visit (RR 0.41 95%CI 0.29 to 0.58), reduce ICU admission (RR 0.47 95%CI 0.23 to 0.92) and mortality (RR 0.32 95%CI 0.13 to 0.77) from the disease.

*Meaning:* In COVID-19 pandemic, the use of bamlanivimab may be warranted. Combinations of two or more monoclonal antibody increase the effect.

## INTRODUCTION

The Coronavirus disease 2019 (COVID-19) has become a major public hazard caused by severe acute respiratory syndrome coronavirus 2 (SARS-CoV-2). By August 2021, more than 203 million people have been reported to be infected with COVID-19, and 4 million people have died of it. Although the advent of vaccines has played a great role in epidemic prevention and control. In developing countries, the availability of vaccines has been slow^[1]^. Several of these COVID-19 therapeutic candidates are currently being studied in preclinical and clinical research stages around the world. Antibody based on viral spike protein (S protein) appears to be the main target of antibody development candidates^[2]^.

Bamlanivimab, also known as LY-CoV555, is a lgG monoclonal antibody which mediate its effects by means of binding with the receptor-binding domain of SARS-CoV-2 to viral neutralization, complement activation and antibody-dependent cellular cytotoxicity^[3, 4]^. In November 2020, the Food and Drug Administration (FDA) authorized an emergency use authorization (EUA) for bamlanivimab, for treatment of mild to moderate COVID-19 in non-admitted patients at high risk for severe disease^[5]^. The benefits or harms of such monoclonal antibodies is limited. Thus, this meta-analysis, aims to summaries efficacy of bamlanivimab use in COVID-19 based on available evidence and find out the potential association between treatment with bamlanivimab and outcomes of COVID-19.

## METHOD

This meta-analysis was described by the Preferred Reporting Items for Systematic Review and Meta-Analyses (PRISMA-P) statement^[6]^. and the guidelines of the Meta-analysis of Observational Studies in Epidemiology^[7]^. The protocol for this study has been registered in the International Prospective Register of Systematic Reviews (PROSPERO, CRD42021272789).

### Search strategy and study selection

Two experience researchers (Bei He and Xuan Cheng) based on The Peer Review of Electronic Search Strategies checklist^[8]^ searched Web of Science, PubMed, Embase, the Cochrane Library, MedRxiv for clinical trials and observational studies. A combination of search terminologies (“Coronavirus Disease 2019” OR “COVID-19” OR “SARS-Cov-2” OR “2019-nCoV Diseases” OR “COVID 19 Virus Infection”) AND (“LY-CoV555” OR “bamlanivimab”). Search was down on August 5, 2021 which was imposed restriction on the date between 1.1 2020 to 8.5 2021. No restriction on the geographical location or language of the studies. To be eligible for inclusion, studies must divide patients into treatment group using bamlanivimab and control group with no bamlanivimab. We also required that the studies reported data on any of the pre-determined outcomes. The studies aimed at preventing the occurrence of COVID-19 were excluded.

### Data extraction

Two reviewers (Huai-rong Xiang, Yun Li) independently screened the titles and abstracts of the studies to retrieve articles and extracted data based on the inclusion and exclusion criteria. Any discrepancies will be resolved through discussion (with a third author, if necessary).

Each included article was thoroughly reviewed, the following data and information were extracted:

(1) The first author’s name, publication date, country and study type.
(2) Patients’ information
(2) Treatment plan (including dosage, timing of using bamlanivimab).
(3) Outcome indicators.

Data was extracted and entered to a pre-defined and piloted Microsoft excel database.

### Quality assessment

Two reviewers (Qi-zhi Zhang and Huai-rong Xiang) independently evaluated the quality of these literature. In case of any disagreement, the third reviewer (Yun Li) consulted for reconciling any difference of opinion. The version 2 Cochrane Risk of bias Assessment tool^[9]^ was used to evaluate the methodological quality of the RCTs over five domains of selection, performance, blinding, completeness, and reporting. For each domain, risk of bias judgements is ‘high’, ‘unclear’ or ‘low’. The Newcastle-Ottawa Scale (NOS)^[10]^ was to evaluate the quality of the cohort studies which comprises three parts of patient selection, comparability, and research results. The total score is 9 which was divided into three categories: (a) high risk (1-3); (b) some concerns (4-6); (c) low risk (7-9).

### Outcomes and subgroup analyses

Primary outcomes were subsequent hospitalization or emergency department (ED) visit, all-cause mortality. Secondary outcomes were admission to intensive care unit admissions. We conducted a priori determined subgroup analyses to explore the source of heterogeneity among observational studies. We performed stratified analyses by (1) the dose of bamlanivimab (700mg, 2800mg, 7000mg and combination) (2) 14 days or 28 days outcomes.

### Statistical Analysis

All statistical analysis was conducted by Stata 16 (StataCorp, College Station, SE) software. Statistical heterogeneity was evaluated by the I-squared (I^2^) test. A fixed-effect model was selected if I^2^ < 40%. When I^2^ value > 40%, heterogeneity was considered significant, and the random-effect model was applied. We used relative risk (RR) for dichotomous data with 95% confidence interval (CI). Publication bias was detected using Egger’s test.

### Certainty of evidence

The certainty of evidence was rated using the Grading of Recommendations, Assessment, Development and Evaluations (GRADE) approach.

## RESULTS

### Search results and study characteristics

As shown in Figure. 1, the total search process yielded 460 records. Following removing duplicates publications 243 studies were remained. After screening of titles and abstracts we excluded 156 studies as these include reviews, commentaries, mechanism researches, and irrelevant to COVID-19. After comprehensively screening 77 full texts and assessment of risk of bias, only 10 eligible studies were included, 3 RCTs^[11-13]^ and 7 observational studies^[14-20]^. We considered observational studies with an NOS score ≥6 with high quality. (Figure 1) All studies included were conducted in America. Assessment of bias for the 9 observational studies revealed a moderate risk of bias as assessed against various quality parameters. (Supplementary table 1). 3 RCTs were classified as low risk because they have been clinically registered in USA National Institutes of Health Register (ClinicalTrials.gov) and had registration numbers. (Supplementary table 2). Two RCTs^[11, 12]^ shared one registration numbers (NCT04427501), but the patients were enrolled at totally different time. One RCT^[11]^ has four bamlanivimab arm for the dose. Owing to the different dosage and limited studies, we conducted a subgroup analysis based on the infusion time at 700 mg bamlanivimab.

**Figure 1:**
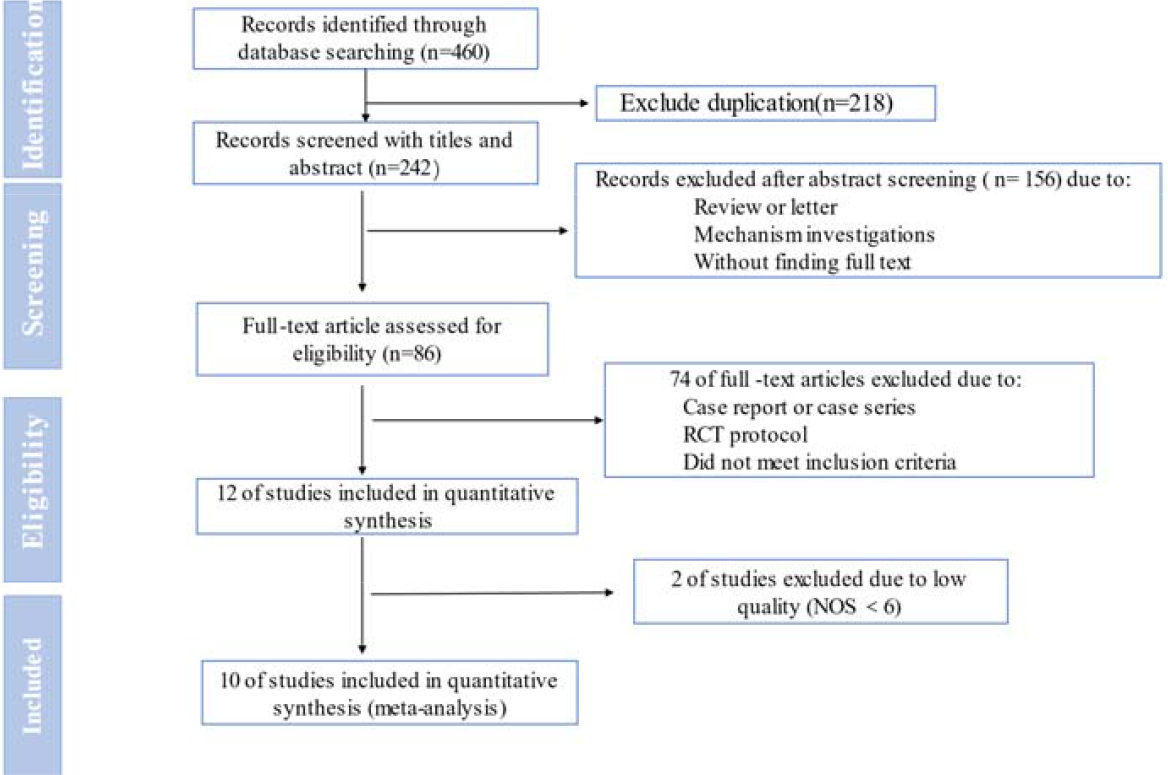
Preferred Reporting Items for Systematic Review and Meta- analysis (PRISMA) flow diagram for the systematic review and meta- analysis.

### Outcome of bamlanivimab for COVID-19

#### Hospitalization or ED visit

Eleven trials (n = 14152 participants) reported on hospitalization or emergency department visit. The use of bamlanivimab compared to placebo could reduce the risk of hospitalizations or emergency department visit (RR 0.41, 95% CI 0.29 to 0.58; *I*^2^= 77.3%, P = 0.000) with low certainty of evidence. (Figure 2) Sensitivity analysis after deleting the study successively revealed the same high heterogeneity.

**Figure 2:**
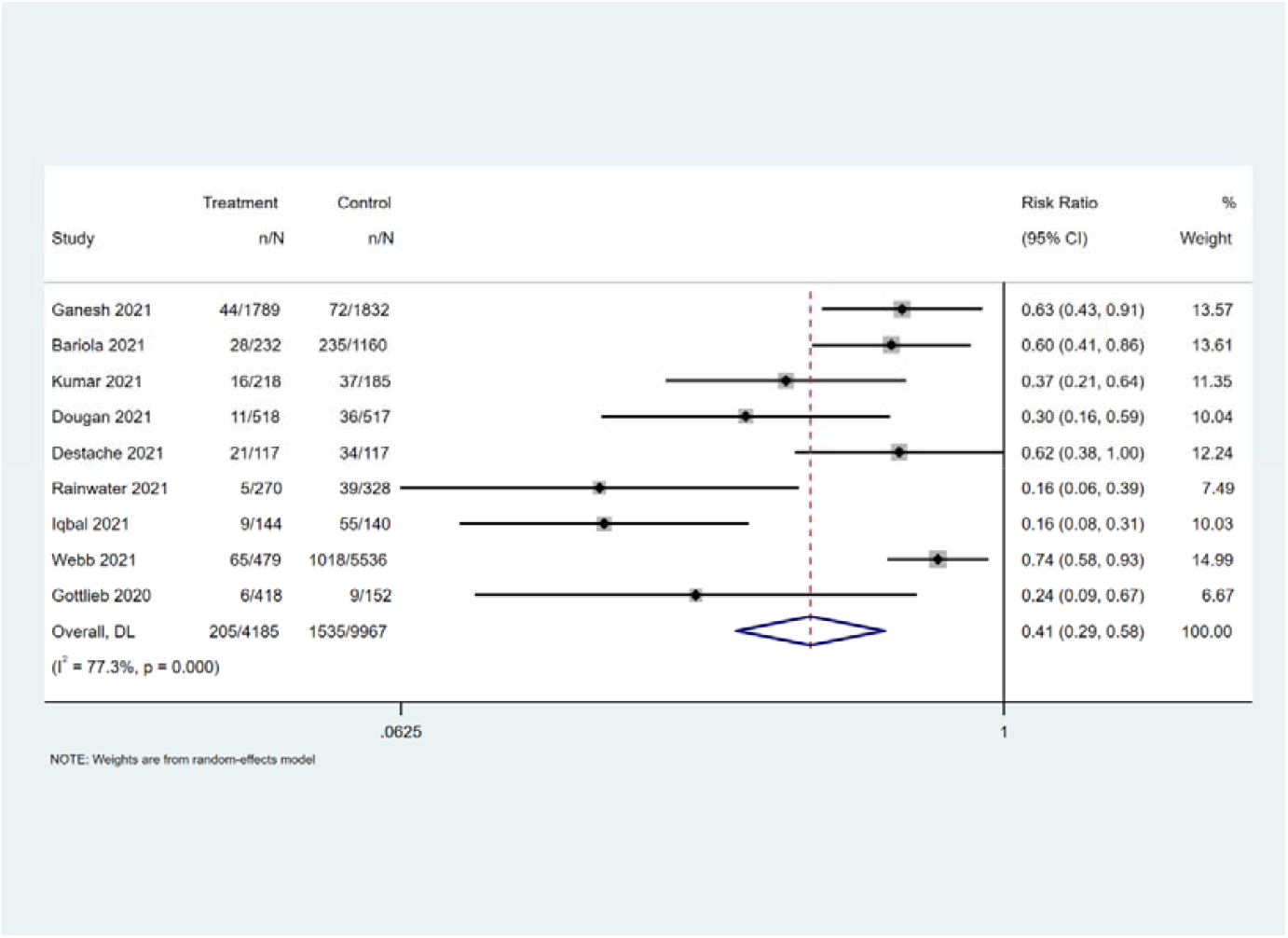
Forest plot of pooled hospitalization or ED visit rate across all included studies. Weights are from random-effects analysis. Note: CI = Confidence Interval, ED = Emergency Department

#### Mortality

Nine trials (n = 13298 participants) reported on mortality. The use of bamlanivimab compared to placebo could reduce the risk of mortality (RR 0.32, 95% CI 0.13 to 0.77; *I*^2^= 47.5%, P = 0.064 low certain of evidence). (Figure 3) We conducted a sensitivity analysis to investigate the source of the heterogeneity. The study conducted by Lundgren et al. included hospitalized patients and the heterogeneity may be caused by different degrees of severity of the disease in the participant. After excluding that study, the heterogeneity decreased significantly (*I*^2^= 9.4%), and the results showed that the bamlanivimab could significantly decrease the rate of mortality. (RR 0.2 95%CI 0.10 to 0.40)

**Figure 3:**
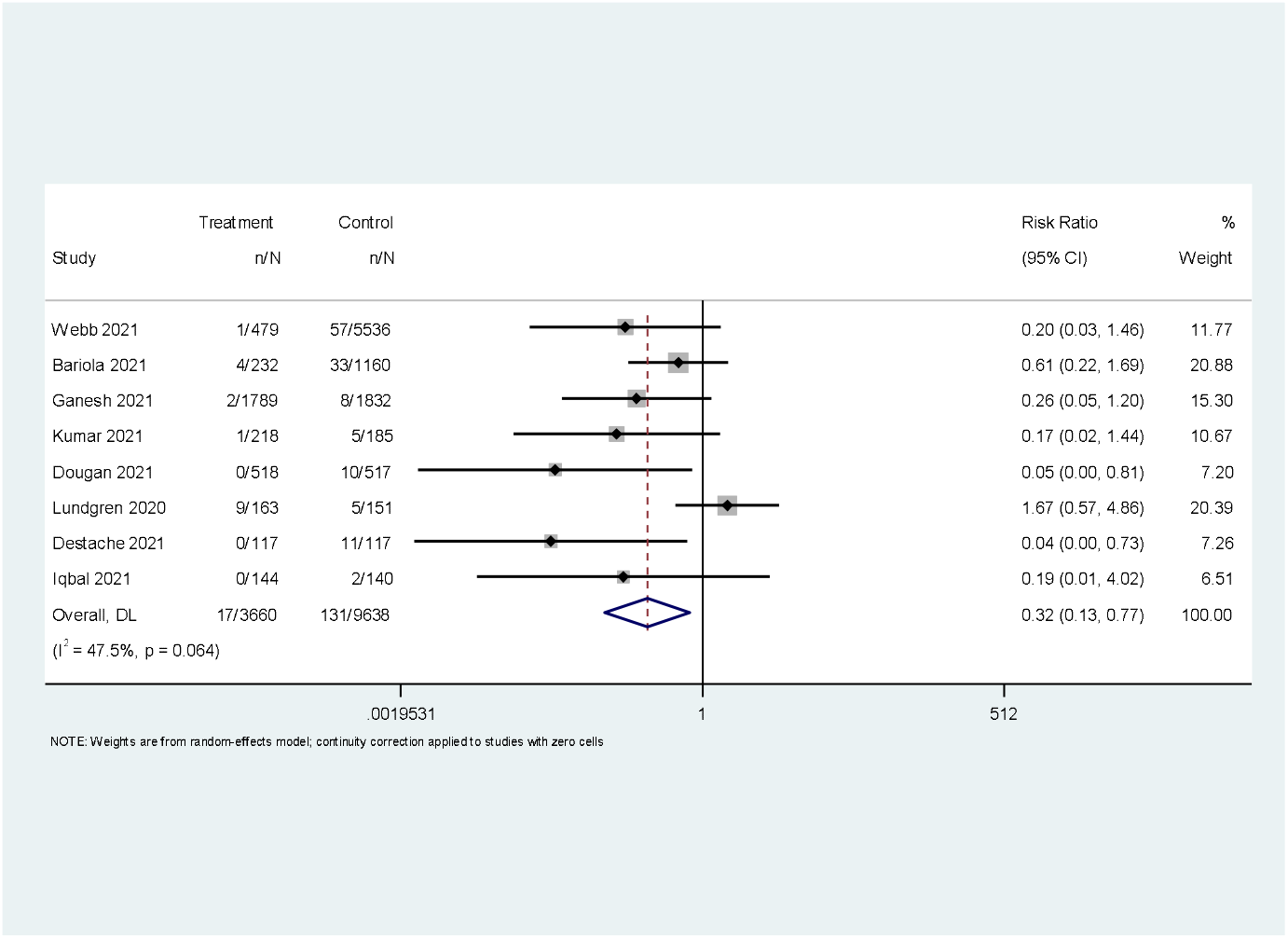
Forest plot of pooled mortality rate across all included studies. Weights are from random-effects analysis. Note: CI = Confidence Interval.

#### ICU admission

Only two trials (n = 5073 participants) reported on mortality. The use of bamlanivimab compared to placebo could reduce the risk of ICU admission (RR 0.47, 95% CI 0.23 to 0.92; *I*^2^= 0.0%, P = 0.672 low certainty of evidence). (Figure 4) We did not perform sensitivity analysis.

**Figure 4:**
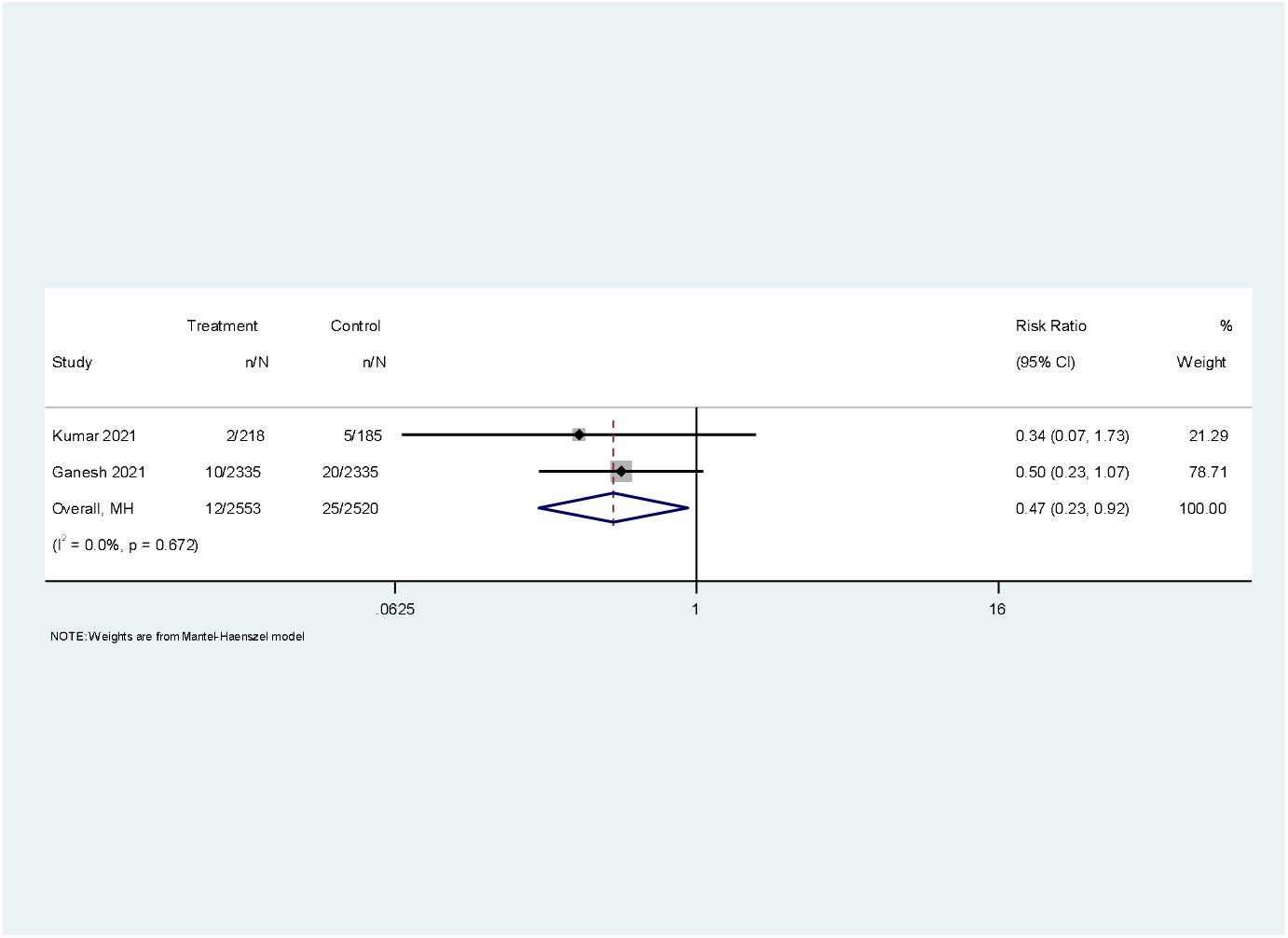
Forest plot of pooled ICU admission rate across all included studies. Weights are from random-effects analysis. Note: CI = Confidence Interval.

#### Safety

In this meta-analysis, 3 RCTs^[11-13]^ reported adverse effects. The pooled result from three trials (n = 1873 participants) found bamlanivimab increased the risk of serious adverse events but no statistic difference. (RR 1.33 95% CI 0.84 to 2.10; *I*^2^ = 0%, P = 0.783 high certainty of evidence) (Figure 5) We did not perform sensitivity analysis about this result.

**Figure 5:**
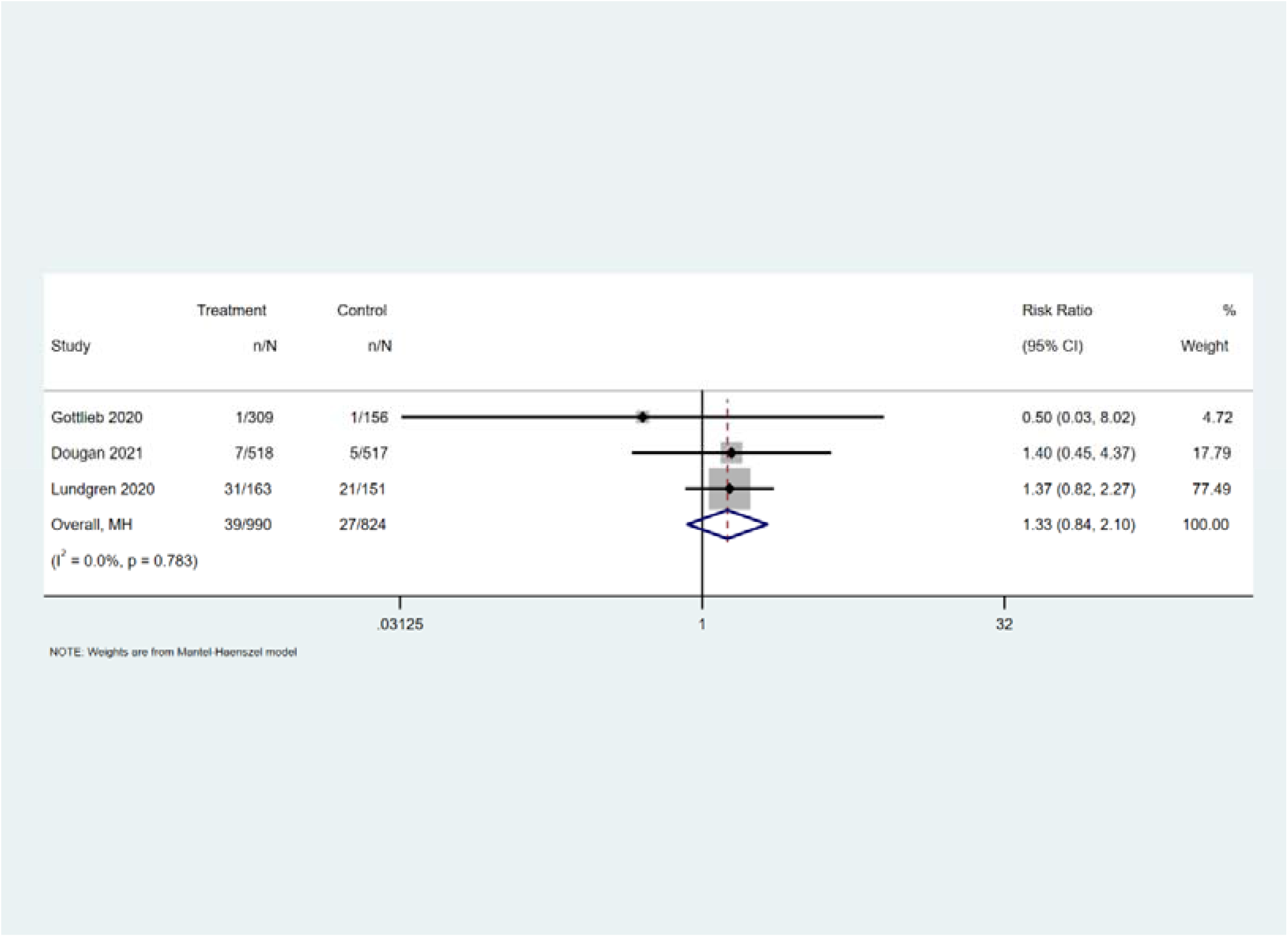
Forest plot of pooled serious adverse events rate across all included studies. Weights are from Fixed-effects analysis. Note: CI = Confidence Interval.

### Subgroup Analysis

#### Subgroup analysis based on dose

Subgroup analysis showed that the 700mg single bamlanivimab and 2800mg bamlanivimab plus 2800mg etesevimab could reduce the risk of hospitalizations or ED visit compared with the control group. And the combination of two antibodies could reduce more risk than 700mg single antibody (RR:0.29 VS 0.59). Due to only one study in the 2800mg single dose and 7000mg dose, there are no statistic difference between treatment and control group. (Figure 6)

**Figure 6:**
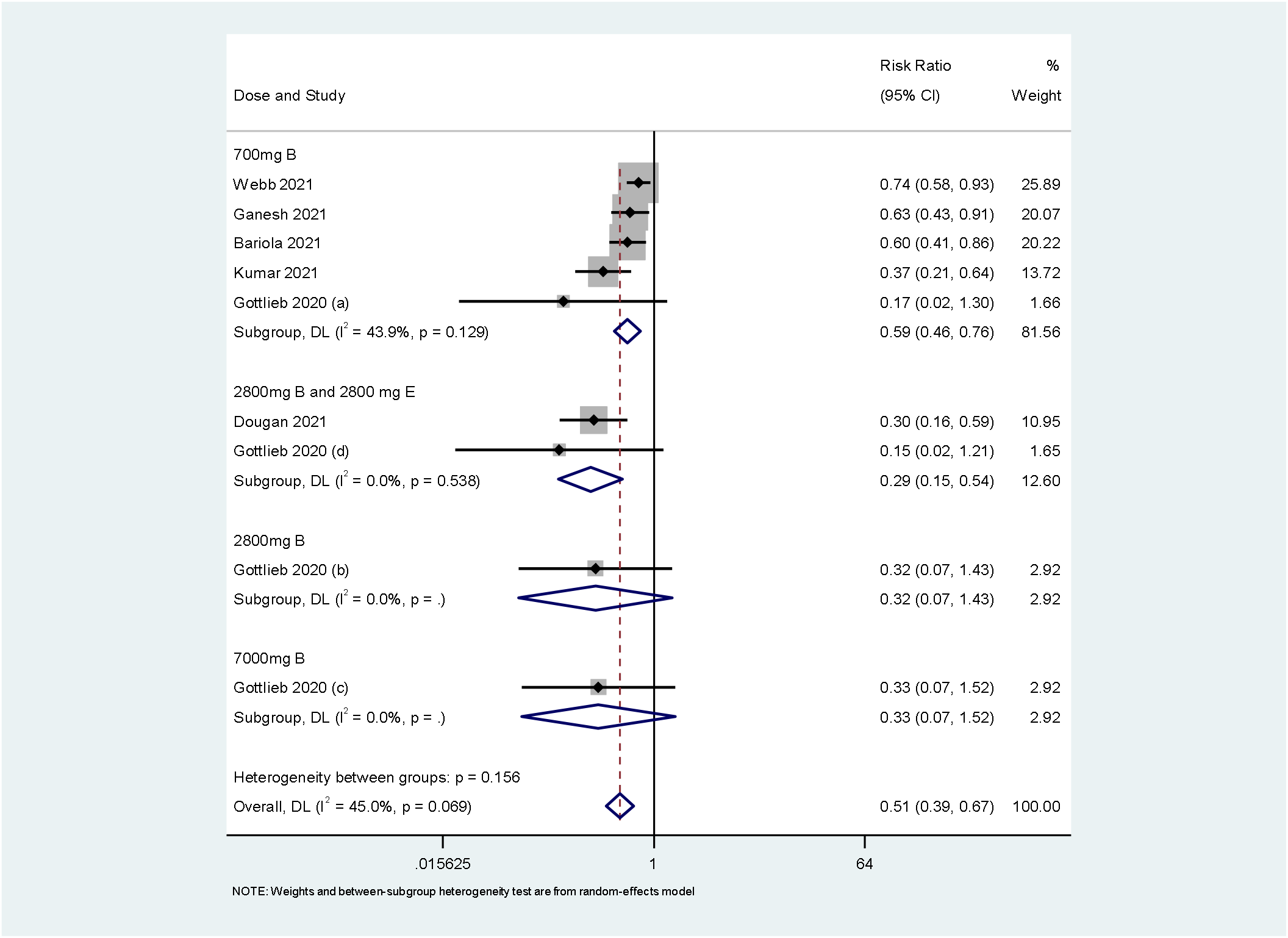
Forest plot for subgroup analysis of hospitalization or ED visit rate by type of dose. Weights are from random-effects analysis. Note: CI = Confidence Interval. 2800mg B and 2800 E = 2800mg bamlanivimab plus 2800mg etesevimab

In the part of mortality, there was no significant difference among 7000mg treatment group and control group. Both 700mg and 2800mg bamlanivimab plus 2800mg etesevimab could decrease the mortality (700mg: RR 0.33 95%CI 0.18 to 0.58; *I*^2^ = 0%, P=0.799. 2800mg bamlanivimab plus 2800mg etesevimab: RR 0.05 95%CI 0.00 to 0.81) (Figure 7)

**Figure 7:**
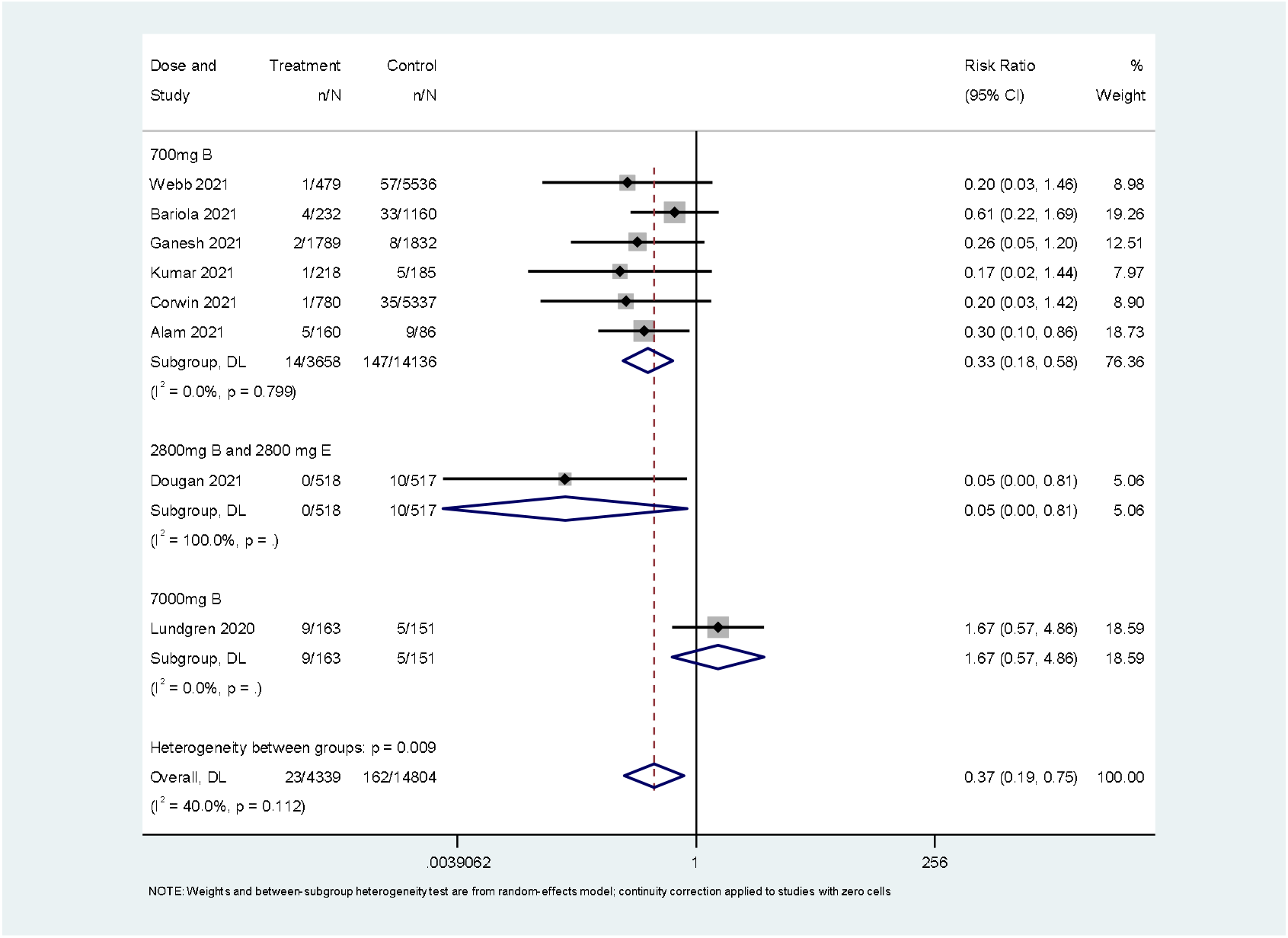
Forest plot for subgroup analysis of mortality rate by type of dose. Weights are from random-effects analysis. Note: CI = Confidence Interval. 2800mg B and 2800 E = 2800mg bamlanivimab plus 2800mg etesevimab

### Risk of publication bias

Assessment of publication bias using Egger’s test showed that there was significant potential publication bias among the included trials hospitalization or ED visit (p=0.001) and mortality (p=0.006). (Supplementary Figure 1)

## DISCUSSION

This is the first meta-analysis included 3 RCTs and 9 observational studies investigating the effect of bamlanivimab with or without etesevimab on clinical outcomes. Despite our inclusion criteria that did not specify the severity of the disease, all the studies were conducted with non-hospitalized patients except the RCT by Lundgren 2020 et al^[13]^. This study provided better evidence for COVID-19 treatment decisions through systematically reviewed, summarized evidence. And our meta-analyses suggested that bamlanivimab may help outpatients to prevent hospitalization, reduce severity and mortality from the disease. What’s more, the combination of bamlanivimab and etesevimab could play a bigger role in the outcomes. Last but also important, early treatment is more effective. There is slightly trend on increasing serious adverse events. The certainty of the evidence of the all-cause hospitalization or ED visit and mortality was low. All observational studies adjust the age confounders, but patients in exposed group had higher risk factor for severe COVID-19 than nonexposed group. And there still was a trend toward a lower risk of hospitalization in bamlanivimab treated patients. Most studies’ patients we included were at mild or moderate severity COVID-19. Only one RCT^[13]^,included hospitalized patients whose infusion time was within 12 days of symptom onset, which announced that no clinical benefit of bamlanivimab administration in patients with already present symptoms of COVID-19. Another study^[21]^ showed the contrary results which only treated to hospitalization. But early intervention of this study is rather comparable to the recently outpatient studies.

It has been proposed that bamlanivimab has ability to neutralize the spike protein of SARS-CoV-2 and consequently block attachment to the human angiotensin-converting enzyme 2 (ACE-2) receptor and deny host cell entry. The FDA announced that bamlanvimab has a definite therapeutic effect on COVID-19. However, Lilly, the company that makes bamlanivimab suggested that it would be unlikely to help patients who admitted to hospital with COVID-19. What’s more, potential drawbacks of monoclonal antibodies have also become clear, such as immune escape by selection of viral mutations^[22]^. For instance, E484K-carrying SARS-CoV-2 variants could resistant to bamlanivimab which emergence and expansion of had been at least partially underestimated^[23]^ And bamlanivimab lost its ability to bind to the spike protein and no longer neutralized the Delta variant^[24, 25]^. Combinations of two or more monoclonal antibodies or polyclonal antisera may therefore increase the barrier sufficiently to prevent escape of the immune system as known from other viral infections^[26]^.

There are some limitations on our meta-analysis. First, groups in some studies were unlikely considered to be comparable on each variable used in the adjustment such as immunosuppression and comorbidity. Second, the small number of participants in some RCTs may increase the likelihood of type II statistical error. Larger scale RCTs are required to confirm the findings. Third, the primary focus of this meta-analyses was cohort studies. Theoretically, observational studies would overestimate the treatment effect of the interventions ^[27]^. Fourth, the data were too limited to perform more meaningful outcome or subgroup analyses. Fifth, the results of egger’s test mean there were high risk of selective reporting or of publication bias.

## Conclusion

Our study showed that bamlanivimab has might be beneficial for mild or moderate ill patients with COVID-19. Combinations of two or more monoclonal antibody increase the effect. Well-designed clinical trials to identify the clinical and biochemical characteristics in COVID-19 patients’ population that could benefit from bamlanivimab are warranted in the future.

## Data Availability

All data relevant to the study are included in the article or uploaded as supplementary information.

## Authors’ Contributions

Wen-xing Peng, and Huai-rong Xiang designed this study; Xuan Cheng and Bei He ran the searched strategy; Huai-rong Xiang and Yun Li selected articles and extracted data; Qi-zhi Zhang and Yun Li evaluate the quality of the literature. Huai-rong Xiang wrote the manuscript, Xuan Cheng, Yun Li, Bei He and Qi-zhi Zhang edited. All listed authors reviewed and approved the final manuscript.

## Data availability statement

All data relevant to the study are included in the article or uploaded as supplementary information. The data are available by accessing the published studies listed in table 1.

**Table 1.**
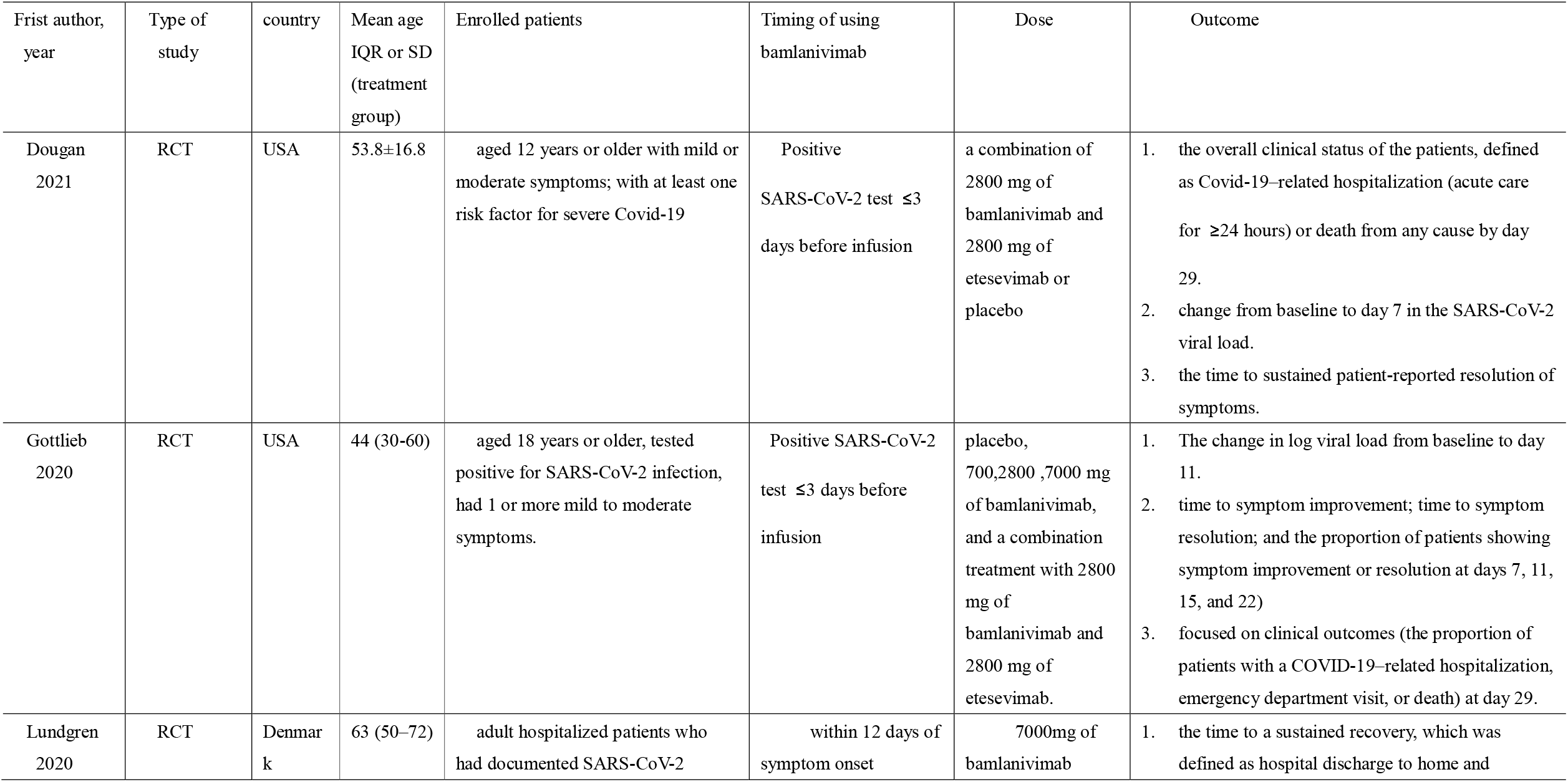
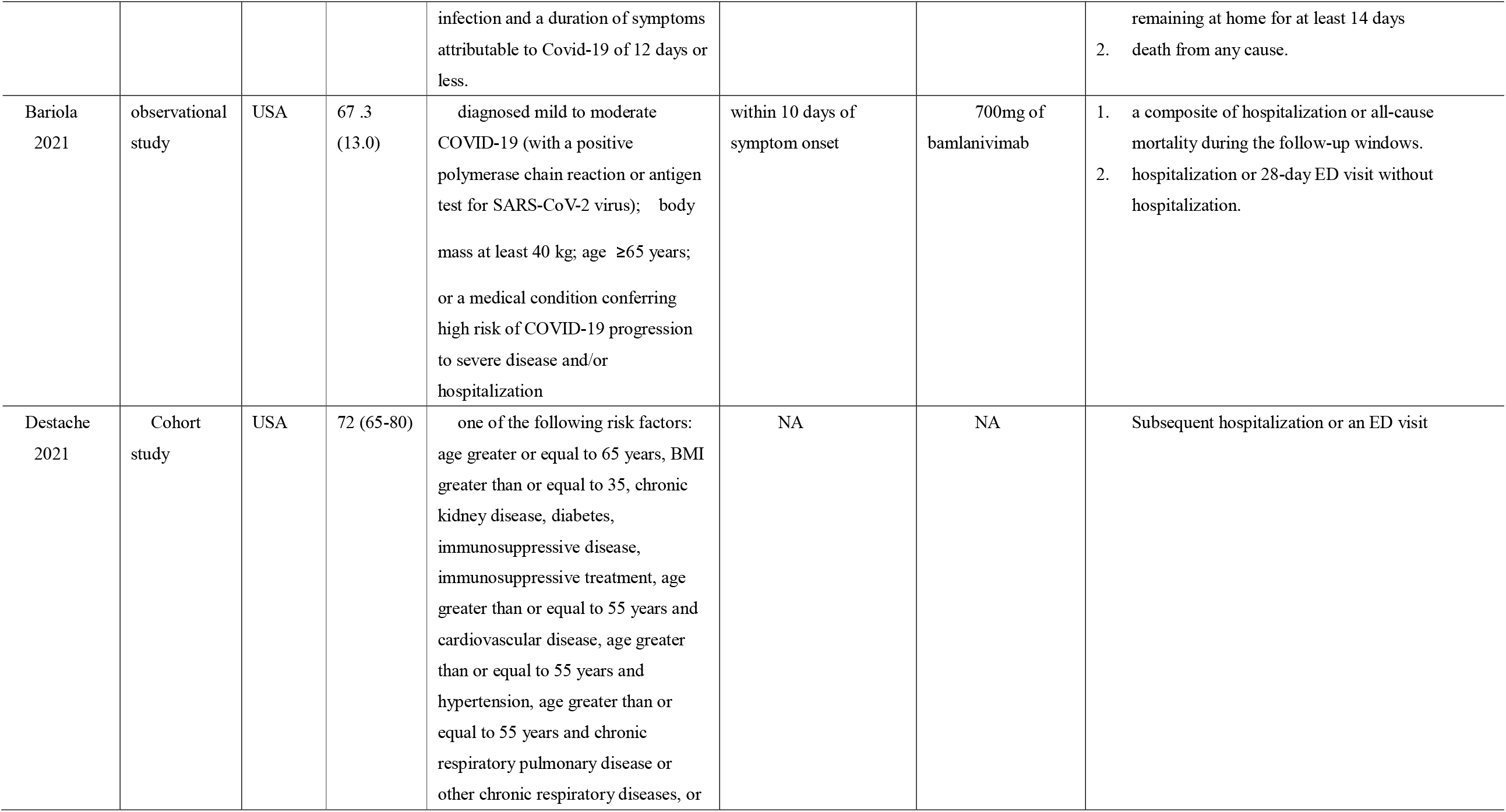
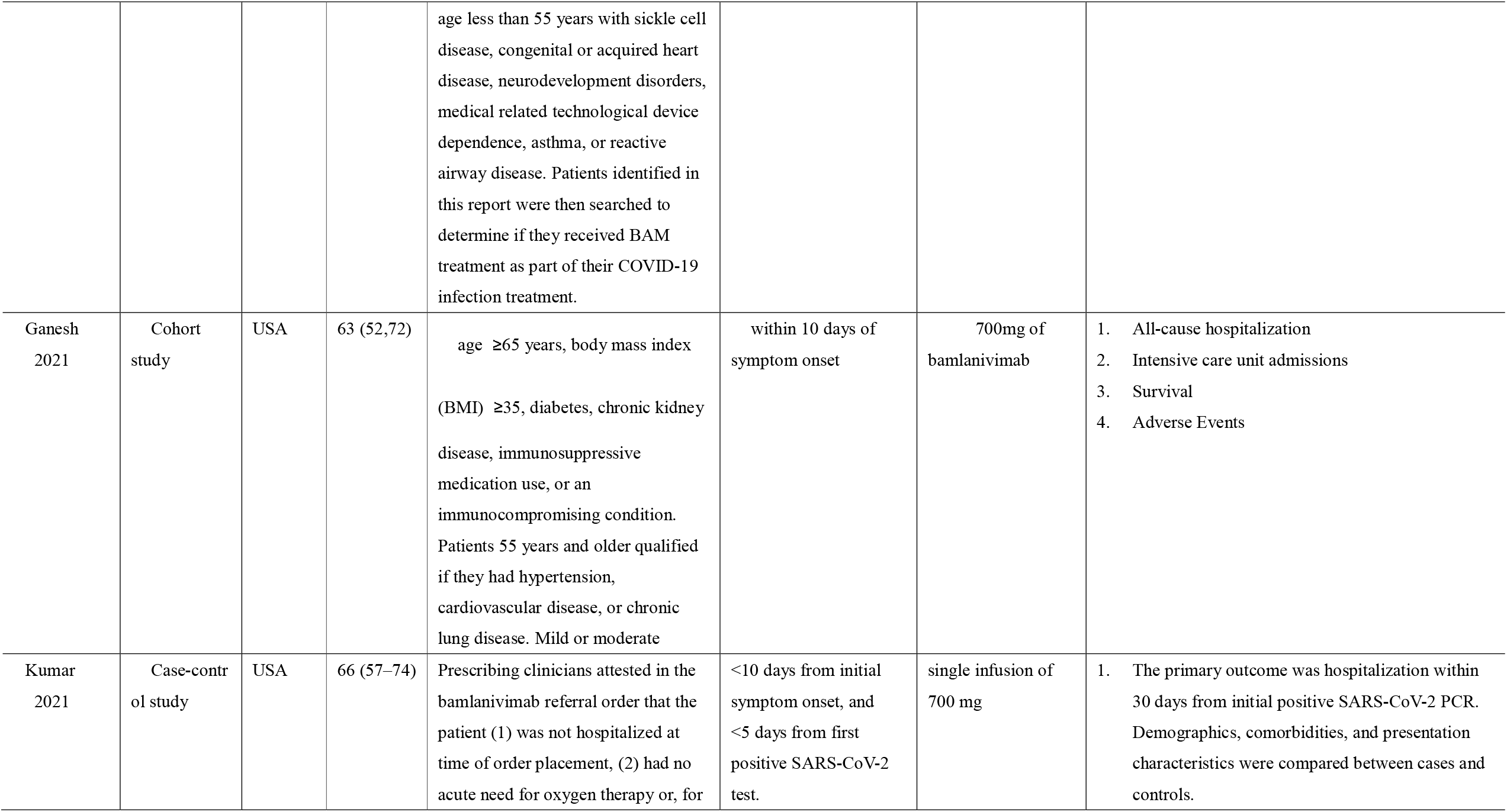
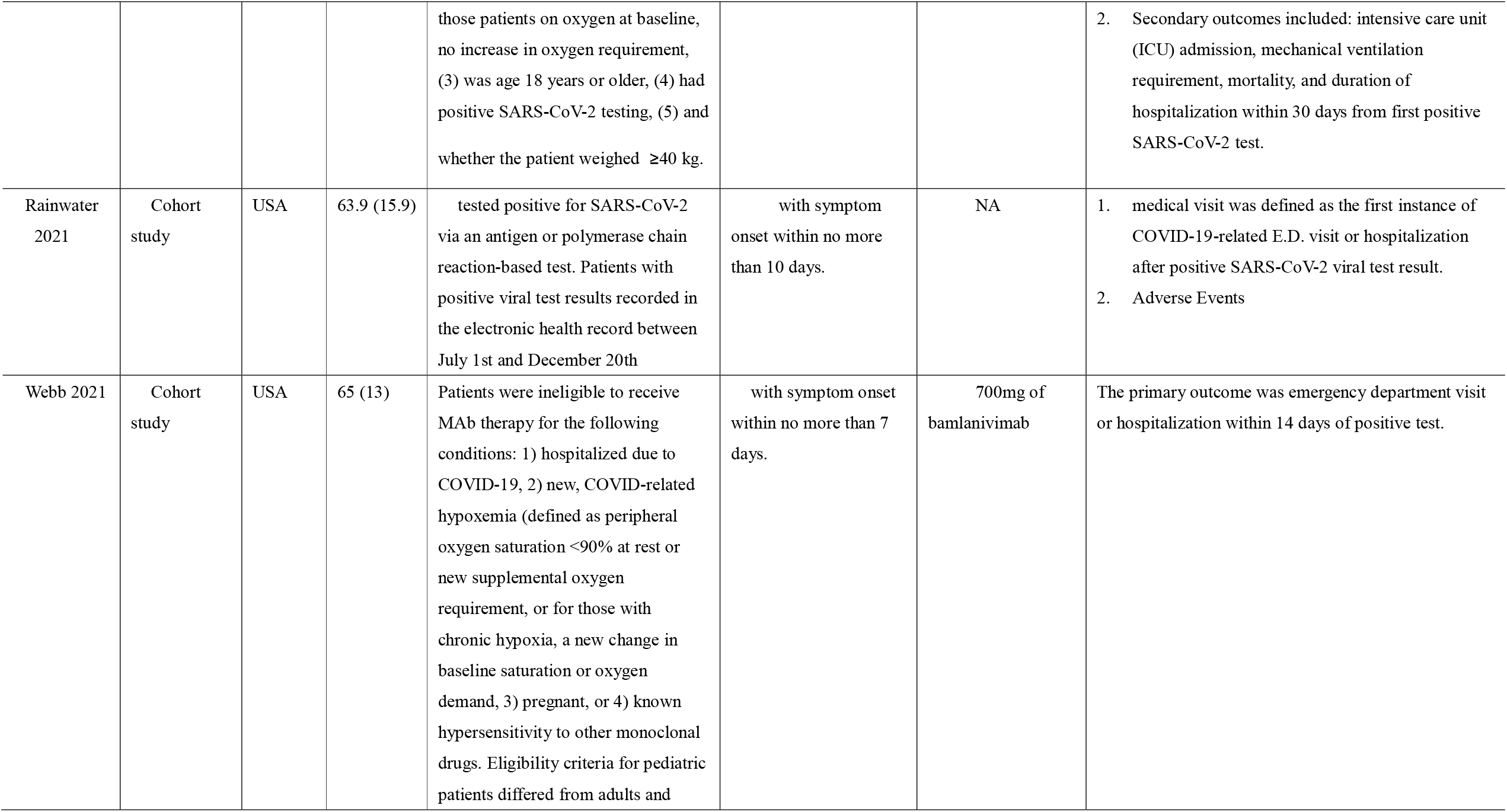
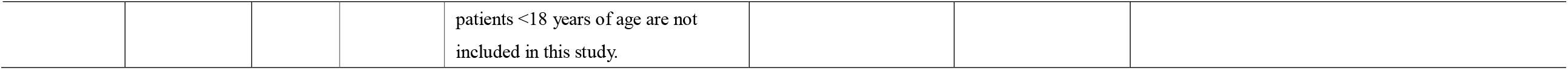
Characteristics of the studies included in the meta□analysis.

| Frist author, year | Type of study | country | Mean age IQR or SD (treatment group) | Enrolled patients | Timing of using bamlanivimab | Dose | Outcome |
| --- | --- | --- | --- | --- | --- | --- | --- |
| Dougan 2021 | RCT | USA | 53.8±16.8 | aged 12 years or older with mild or moderate symptoms; with at least one risk factor for severe Covid-19 | Positive SARS-CoV-2 test ≤3 days before infusion | a combination of 2800 mg of bamlanivimab and 2800 mg of etesevimab or placebo | the overall clinical status of the patients, defined as Covid-19–related hospitalization (acute care for ≥24 hours) or death from any cause by day 29.<br>change from baseline to day 7 in the SARS-CoV-2 viral load.<br>the time to sustained patient-reported resolution of symptoms. |
| Gottlieb 2020 | RCT | USA | 44 (30-60) | aged 18 years or older, tested positive for SARS-CoV-2 infection, had 1 or more mild to moderate symptoms. | Positive SARS-CoV-2 test ≤3 days before infusion | placebo, 700,2800 ,7000 mg of bamlanivimab, and a combination treatment with 2800 mg of bamlanivimab and 2800 mg of etesevimab. | The change in log viral load from baseline to day 11.<br>time to symptom improvement; time to symptom resolution; and the proportion of patients showing symptom improvement or resolution at days 7, 11, 15, and 22)<br>focused on clinical outcomes (the proportion of patients with a COVID-19–related hospitalization, emergency department visit, or death) at day 29. |
| Lundgren 2020 | RCT | Denmark | 63 (50–72) | adult hospitalized patients who had documented SARS-CoV-2 | within 12 days of symptom onset | 7000mg of bamlanivimab | the time to a sustained recovery, which was defined as hospital discharge to home and |
|  |  |  |  | infection and a duration of symptoms attributable to Covid-19 of 12 days or less. |  |  | remaining at home for at least 14 days<br>2. death from any cause. |
| Bariola 2021 | observational study | USA | 67 .3 (13.0) | diagnosed mild to moderate COVID-19 (with a positive polymerase chain reaction or antigen test for SARS-CoV-2 virus); body mass at least 40 kg; age $\geq 65$ years; or a medical condition conferring high risk of COVID-19 progression to severe disease and/or hospitalization | within 10 days of symptom onset | 700mg of bamlanivimab | 1. a composite of hospitalization or all-cause mortality during the follow-up windows.<br>2. hospitalization or 28-day ED visit without hospitalization. |
| Destache 2021 | Cohort study | USA | 72 (65-80) | one of the following risk factors: age greater or equal to 65 years, BMI greater than or equal to 35, chronic kidney disease, diabetes, immunosuppressive disease, immunosuppressive treatment, age greater than or equal to 55 years and cardiovascular disease, age greater than or equal to 55 years and hypertension, age greater than or equal to 55 years and chronic respiratory pulmonary disease or other chronic respiratory diseases, or age less than 55 years with sickle cell disease, congenital or acquired heart disease, neurodevelopment disorders, medical related technological device dependence, asthma, or reactive airway disease. Patients identified in this report were then searched to determine if they received BAM treatment as part of their COVID-19 infection treatment. | NA | NA | Subsequent hospitalization or an ED visit |
| Ganesh 2021 | Cohort study | USA | 63 (52,72) | age $\geq 65$ years, body mass index (BMI) $\geq 35$, diabetes, chronic kidney disease, immunosuppressive medication use, or an immunocompromising condition. Patients 55 years and older qualified if they had hypertension, cardiovascular disease, or chronic lung disease. Mild or moderate | within 10 days of symptom onset | 700mg of bamlanivimab | 1. All-cause hospitalization<br>2. Intensive care unit admissions<br>3. Survival<br>4. Adverse Events |
| Kumar 2021 | Case-control study | USA | 66 (57–74) | Prescribing clinicians attested in the bamlanivimab referral order that the patient (1) was not hospitalized at time of order placement, (2) had no acute need for oxygen therapy or, for | <10 days from initial symptom onset, and <5 days from first positive SARS-CoV-2 test. | single infusion of 700 mg | 1. The primary outcome was hospitalization within 30 days from initial positive SARS-CoV-2 PCR. Demographics, comorbidities, and presentation characteristics were compared between cases and controls. |
| | | | | those patients on oxygen at baseline, no increase in oxygen requirement, (3) was age 18 years or older, (4) had positive SARS-CoV-2 testing, (5) and whether the patient weighed $\geq 40$ kg. | | | 2. Secondary outcomes included: intensive care unit (ICU) admission, mechanical ventilation requirement, mortality, and duration of hospitalization within 30 days from first positive SARS-CoV-2 test. |
| Rainwater 2021 | Cohort study | USA | 63.9 (15.9) | tested positive for SARS-CoV-2 via an antigen or polymerase chain reaction-based test. Patients with positive viral test results recorded in the electronic health record between July 1st and December 20th | with symptom onset within no more than 10 days. | NA | 1. medical visit was defined as the first instance of COVID-19-related E.D. visit or hospitalization after positive SARS-CoV-2 viral test result.<br>2. Adverse Events |
| Webb 2021 | Cohort study | USA | 65 (13) | Patients were ineligible to receive MAb therapy for the following conditions: 1) hospitalized due to COVID-19, 2) new, COVID-related hypoxemia (defined as peripheral oxygen saturation $<90\%$ at rest or new supplemental oxygen requirement, or for those with chronic hypoxia, a new change in baseline saturation or oxygen demand, 3) pregnant, or 4) known hypersensitivity to other monoclonal drugs. Eligibility criteria for pediatric patients differed from adults and | with symptom onset within no more than 7 days. | 700mg of bamlanivimab | The primary outcome was emergency department visit or hospitalization within 14 days of positive test. |

|  |  |  |  |  |
| --- | --- | --- | --- | --- |
|  |  |  |  | patients <18 years of age are not included in this study. |

**Table 2.**
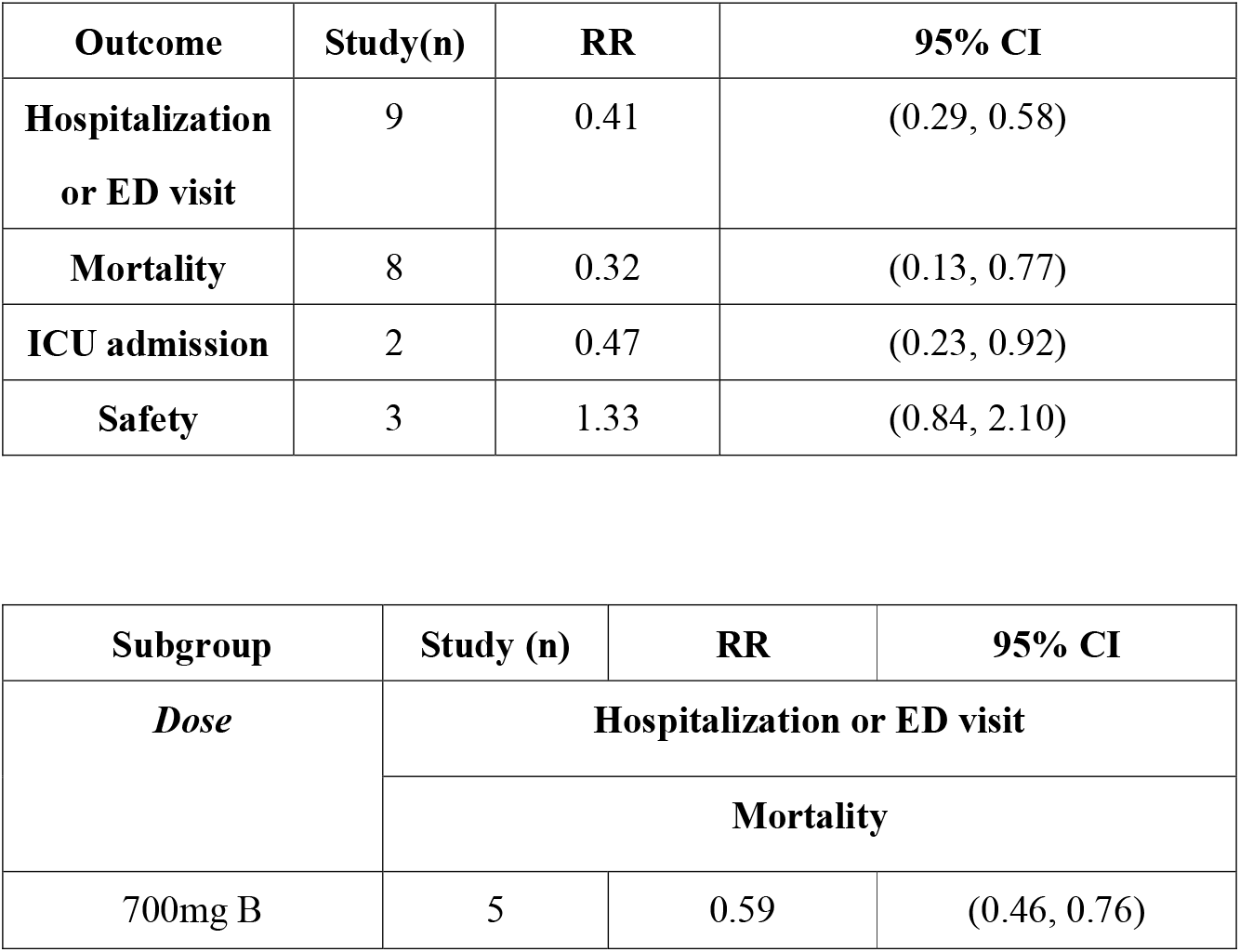
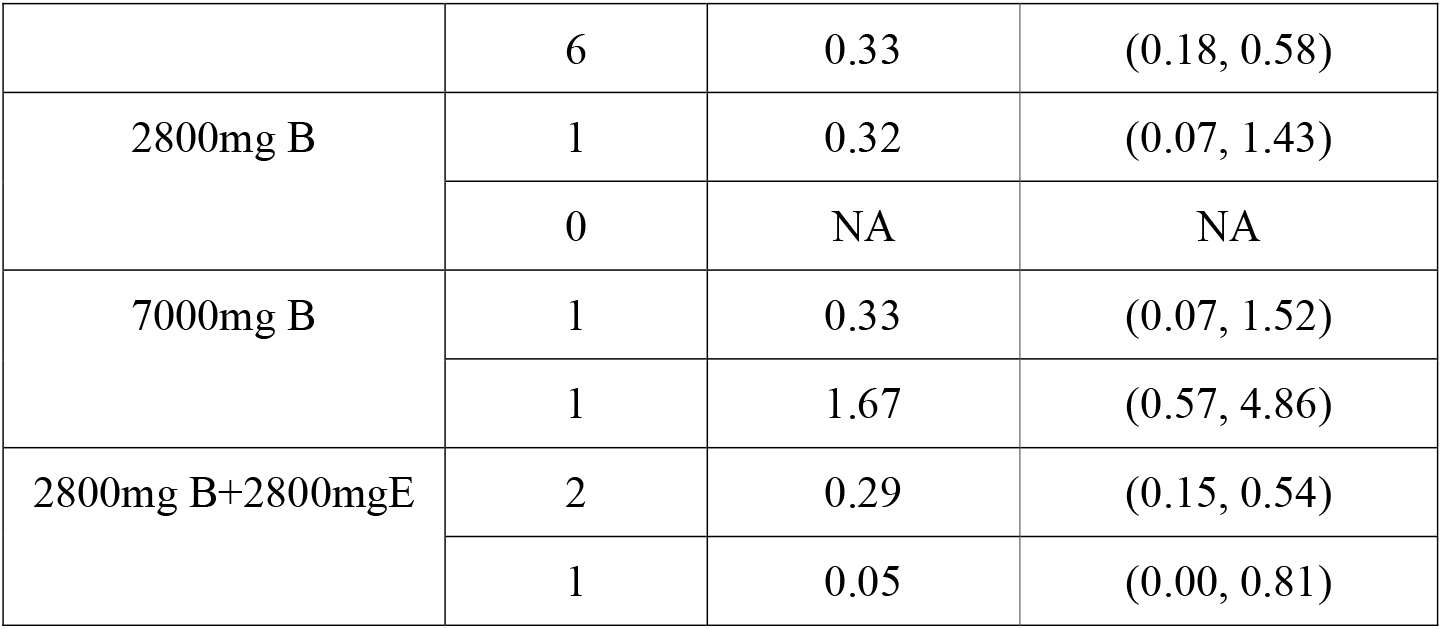
Outcome summary.

| **Outcome** | **Study(n)** | **RR** | **95% CI** |
| --- | --- | --- | --- |
| **Hospitalization**<br>**or ED visit** | 9 | 0.41 | (0.29, 0.58) |
| **Mortality** | 8 | 0.32 | (0.13, 0.77) |
| **ICU admission** | 2 | 0.47 | (0.23, 0.92) |
| **Safety** | 3 | 1.33 | (0.84, 2.10) |

| **Subgroup** | **Study (n)** | **RR** | **95% CI** |
| --- | --- | --- | --- |
| ***Dose*** | **Hospitalization or ED visit** |  |  |
|  | **Mortality** |  |  |
| 700mg B | 5 | 0.59 | (0.46, 0.76) |
|  | 6 | 0.33 | (0.18, 0.58) |
| 2800mg B | 1 | 0.32 | (0.07, 1.43) |
|  | 0 | NA | NA |
| 7000mg B | 1 | 0.33 | (0.07, 1.52) |
|  | 1 | 1.67 | (0.57, 4.86) |
| 2800mg B+2800mgE | 2 | 0.29 | (0.15, 0.54) |
|  | 1 | 0.05 | (0.00, 0.81) |

## Acknowledgments

None

---

**Figure 1:**
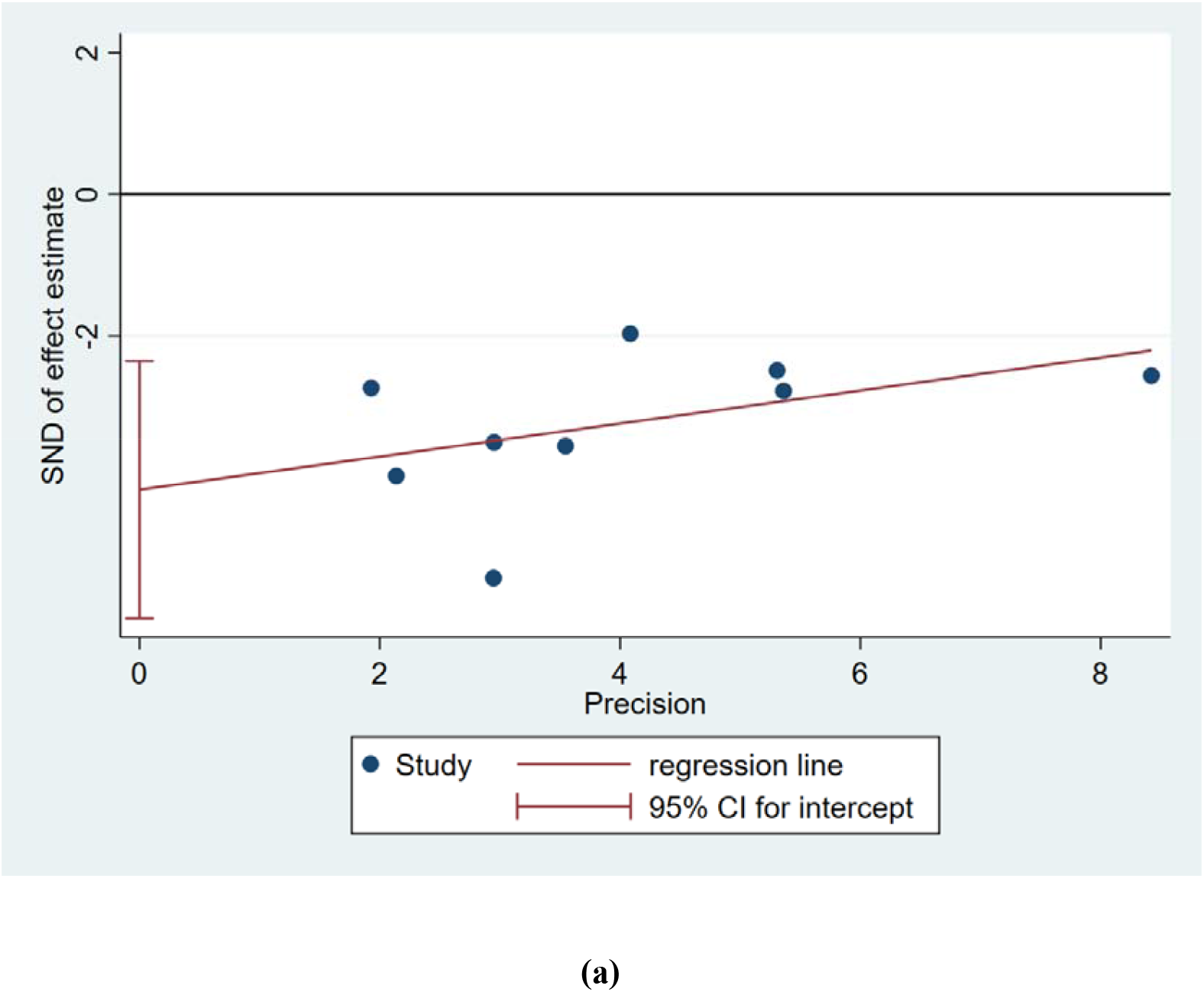
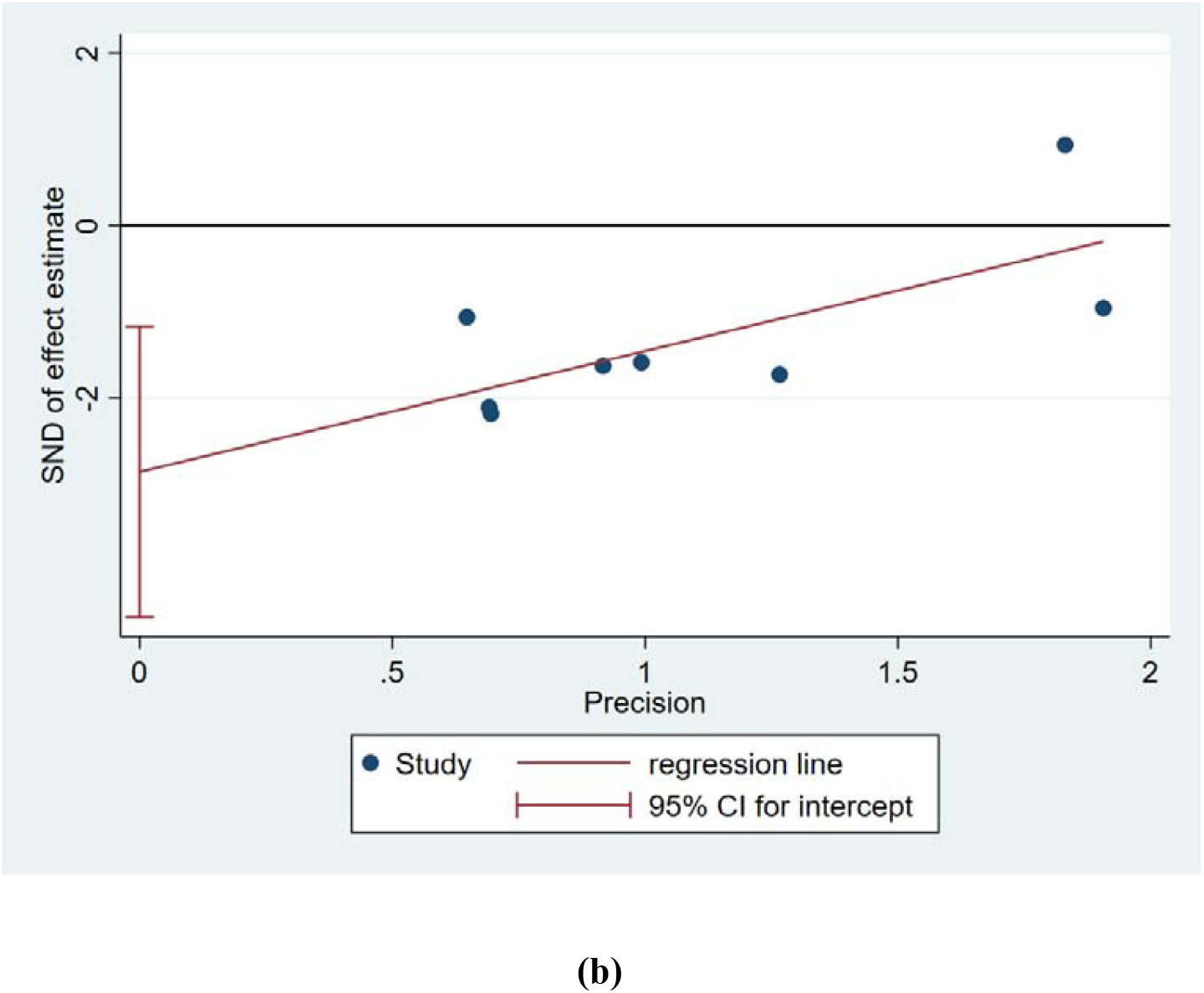
Risk of publication bias. (a)hospitalization or ED visit; (b)mortality.

**Table 1.** Risk of bias summary: the review authors’ judgments about each risk-of-bias item for each included cohort study.

| Study | Patients' selection | Comparability between groups | Outcome or exposure factors assessment | NOS score | Risk of bias |
| --- | --- | --- | --- | --- | --- |
| Bariola 2021 | 3 | 2 | 2 | 7 | L |
| Destache 2021 | 4 | 2 | 2 | 8 | L |
| Iqbal 2021 | 3 | 2 | 3 | 8 | L |
| Ganesh 2021 | 3 | 2 | 3 | 7 | L |
| Kumar 2021 | 3 | 1 | 2 | 6 | M |
| Rainwater 2021 | 3 | 1 | 2 | 6 | M |
| Webb 2021 | 3 | 2 | 1 | 6 | M |

NOS: New-Castle Ottawa scale; The summary risk of bias was determined as low (NOS scores ≥ 7), moderate ( $4\leq$ NOS scores ≤ 6), and high (NOS scores ≤ 3).
L: low risk; M: moderate risk; H: high risk

**Table 2:** Risk of bias summary: the review authors’ judgments about each risk-of-bias item for each included randomized control trial.

| Study | Random sequence generation | Allocation concealment | Blinding of participants and personnel | Blinding of outcome assessment | Incomplete outcome data | Selective reporting | Other bias |
| --- | --- | --- | --- | --- | --- | --- | --- |
| Gottlieb et al (2021) | L | L | L | U | L | L | L |
| Lundgren et al(2021) | L | L | L | L | L | L | L |
| Dougan et al (2021) | L | U | L | U | L | L | L |

L: low risk; U: unclear risk; H: high risk

**Table 3:**
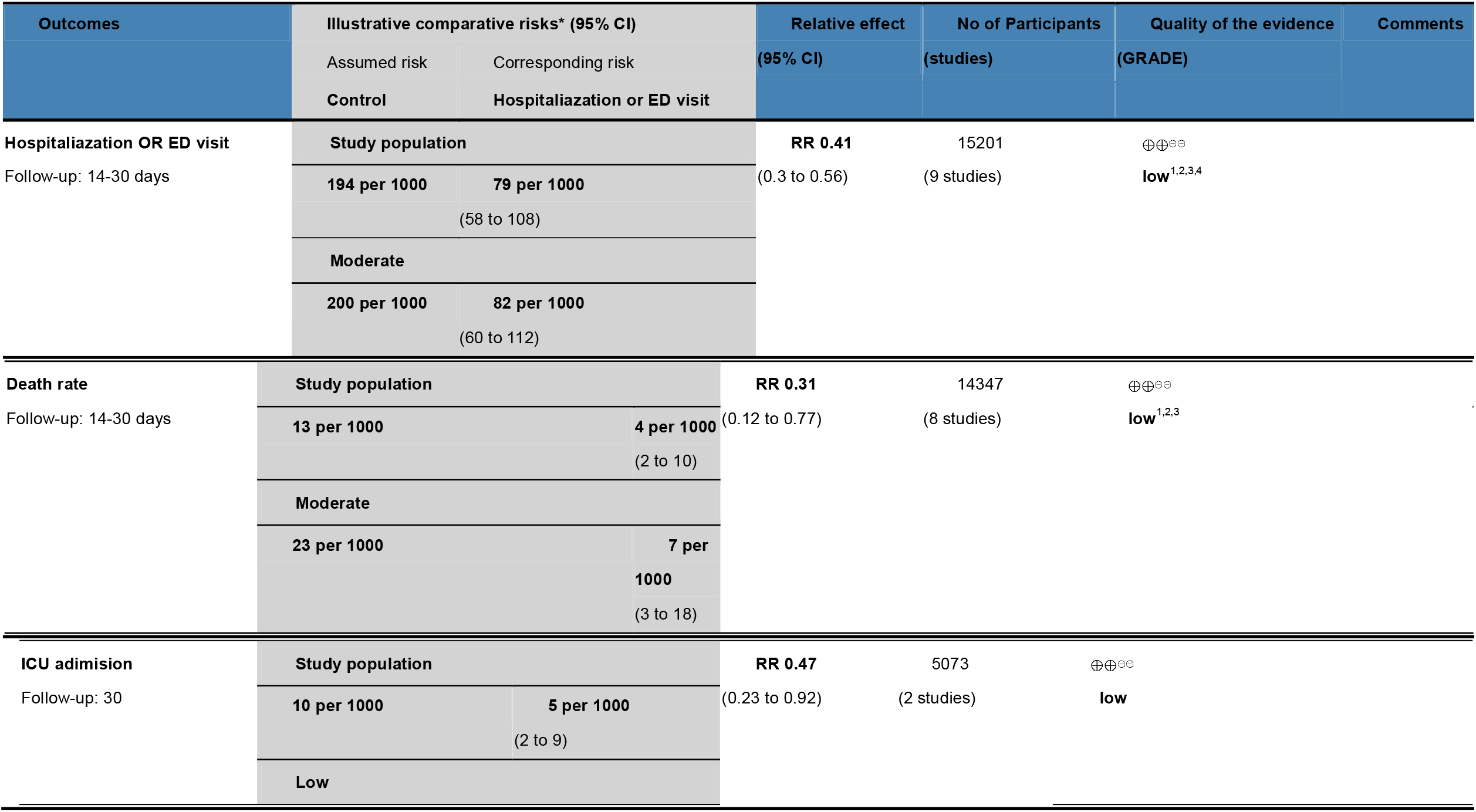
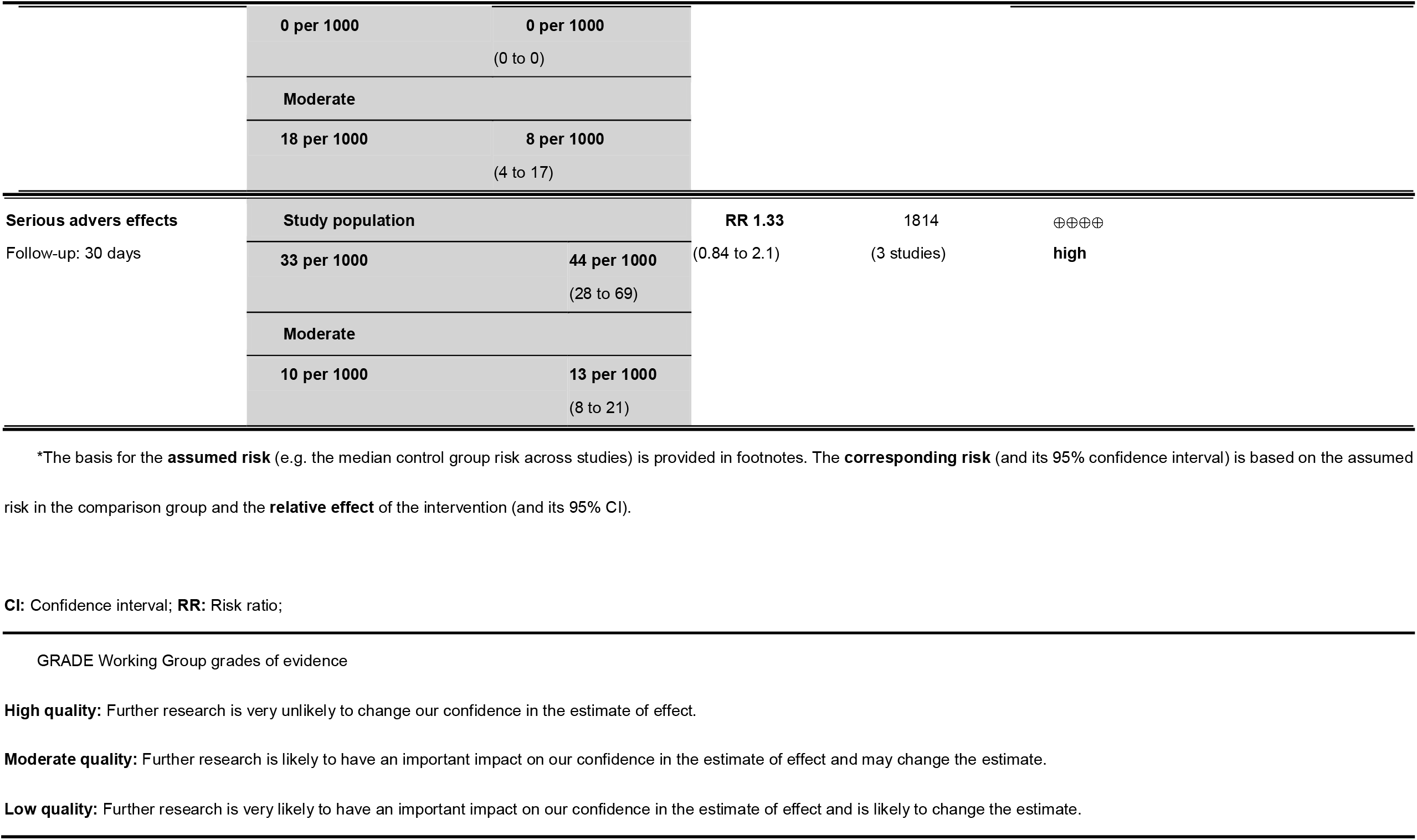
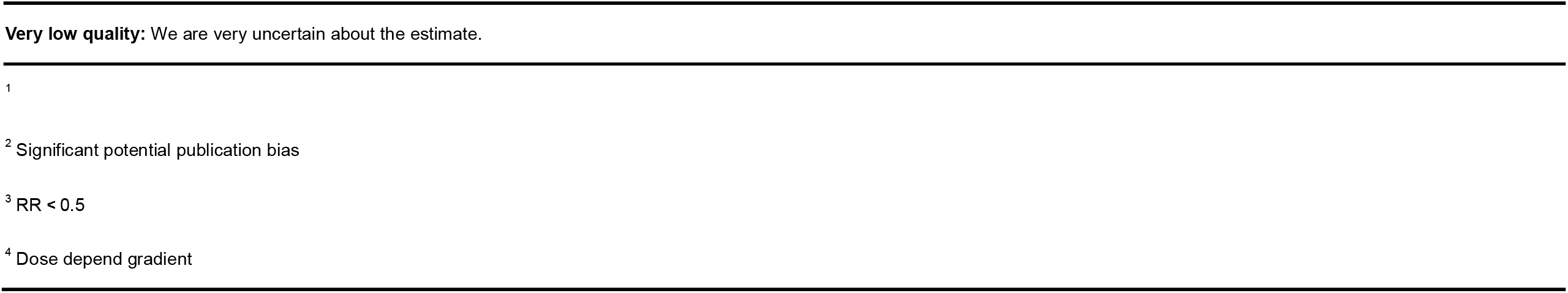
GRADE summary of findings on use of intravenous immunoglobulin in COVID-19.

